# National Health Insurance Scheme: A pathway to a sustained access to medicine in Nigeria

**DOI:** 10.1101/2020.06.23.20138164

**Authors:** Nkoli P. Uguru, Udochukwu U. Ogu, Chibuzo C. Uguru, Ogochukwu Ibe

**Author notes:** **Corresponding author:** Ogu Udochukwu Ugochukwu.

## Abstract

**Objective:** The National Health Insurance Scheme (NHIS) has at its aim the need to ensure that every Nigerian has equal access to good quality health care services. So far, only the Formal Sector Social Health Insurance Program (FSSHIP) aspect of the scheme has been fully activated. The question remains, why the delay towards universal coverage?

**Design:** The study was a cross-sectional and mixed method design. Both qualitative and quantitative methods were utilized for the study.

**Setting:** This study was conducted in NHIS accredited facilities in Enugu State.

**Participants:** A sample of 300 enrolees were selected randomly. For the qualitative study, 6 in-depth interviews (IDIs) were conducted face to face with NHIS desk officers across the three tiers of health care represented.

**Results:** The qualitative findings shows that 94.9% of respondents sought medical help. 78.4% of the respondents indicated that the scheme improved their access to care. The qualitative finding found that there was no discrepancy in access among socio-economic groups. NHIS was reported to have improved access to medicine over the years. In the qualitative, majority of the IDI respondents stated that many of the staff in NHIS accredited facilities are not trained on what is expected or required of them with regards to the scheme.

**Conclusion:** A focus on accessibility, affordability and availability for the scheme means that on account of either of the three, all facility categories and their interests must be considered in further planning of the scheme to ensure that things hold up fine.

**Article Summary:** *Strengths and Limitations:*

- The study participants were only from one state.
- The study only focused on NHIS and NHIS accredited facilities.
- NHIS desk officers, hospital directors and admin officers were the focus for IDIs.

## Introduction

Health is the entry point for breaking the vicious circle of ill health, poverty and under-development and transforming it to improved health status, sustainable development and prosperity (1). Countries across the world in this present time view health insurance as a means to ensure access to health care and protecting patients from financial risks (2,3). Many African countries including Nigeria has established their version of health insurance (4).

The National Health Insurance Scheme (NHIS) which was established with decree 35 of 1999 has at its aim the need to ensure that every Nigerian has equal access to good quality health care services (2). The actual implementation of the NHIS commenced in 2002 and was consolidated in 2005 (2,5). Aspects of the scheme are; Formal Sector Social Health Insurance Program (FSSHIP), the Urban Health Self-employed Social Health Insurance Program (USSHIP) and Rural Community Social Health Insurance Program (RCSHIP), this is a community based health insurance model (5). So far, only the FSSHIP aspect of the scheme has been fully activated. The question remains, why the delay towards universal coverage?

Any health insurance program directed or aimed towards the benefit of the public should consider the following, availability, accessibility and affordability (6). Availability in this instance would mean the existence of adequate staff (skills-mix), drugs and equipment. According to Goudge, Gilson, Russell, Gumede and Mills (7), the shortage of health service often means that appropriate care is not available. Global health workforce alliance (GHWA) (8) posits that availability is the adequate supply and appropriate stock of health workers who have the competency and skill set to match the health needs of the population. Accessibility according to GHWA (8) involves the equitable distribution of health workers by taking into account the demographic composition of rural – urban mix and under-developed areas of the population. Affordability, according to Axene (9) is the ability to purchase a good or service. Affordability determines if a person or organization with limited resources is able to make a purchase or has sufficient income to pay for health care costs (9).

A longitudinal study (7) done, in South Africa on affordability, availability and acceptability barriers to health care for the chronically ill, showed that livelihood exhausted from previous illness and death, low income and limited social network prevented initial healthcare consultations. On the other hand, the monthly expenditure from repeated consultation with possibility of referral to other centers and use of ambulance services was observed to be about 60% of income further hampering access (7). Thus in order to improve access to good quality healthcare, more emphasis should be placed on access to the public sector. The focus should be on improving drug supply chains, ambulance services, clinical capacity at public clinics and most importantly addressing the financial constraint faced by the socially disadvantaged. It is also imperative to think through how providers engage with patients in a way that strengthens their therapeutic alliance.

Rekha, Wajid, Radhakrishnan and Matthew (10) measured accessibility index using a three step floating catchment area in a geographical framework. Three variables were considered; attractiveness of health care centre, travel time or distance between the location of the service centre and residence and population demands for health care facilities.

Another study (11), found that respondents who described quality with regards to the ease with which they got care or short waiting time as good, are 3.9 times more likely to have private facilities as their chosen health care providing facility. Also the data collected indicated that cost for service is 2.9 times more likely to predict the use of public health facility as the usual health provider.

Inadequacies in quality of workforce affects improvement in health outcomes. Given that without these three, availability, accessibility and affordability together, healthcare outcomes in Nigeria will continually be poor. Despite the several failures of the Nigerian health care system, some studies suggest that if managed well, the NHIS could be a useful ground for good health care delivery (12). The purpose of this study therefore, is to emphasize the importance of or role played by access (availability, accessibility and affordability) towards the successful implementation of the NHIS (especially towards its universal coverage goals).

## Methods

The study was a cross-sectional and mixed method design. Both qualitative and quantitative methods were utilized for the study.

### Study setting

This study was conducted in NHIS accredited facilities in Enugu State. The state is situated in the south east part of the country with a population of 4,411,100 million with an annual population growth of 3.0 (13). The state has three senatorial zones (Enugu north, east and west) with 17 local governments. In addition, there are 962 health facilities comprising 4 tertiary, 148 secondary (96 private and 52 public) and 774 (492 public and 282 private) primary health facilities. The federal government funds and operates three tertiary health facilities. One tertiary facility is operated by the state (14).

### Data collection methods

A minimum sample size of 274 respondents was calculated using sample size calculation for community survey. 10% was added to accommodate non-responders thus increasing respondents surveyed to 300. Respondents were drawn from the beneficiaries of the National Health Insurance scheme which are federal civil servants in Enugu State. A sample of 300 enrolees were selected randomly. The questionnaires were interviewer administered and they contained questions eliciting information on socio-demographic details, availability of needed medicines, affordability of needed medicines, perception of quality of medicines and patient satisfaction. For the qualitative study, 6 in depth interviews (IDIs) were conducted face to face with NHIS desk officer across the three tiers of health care represented. The IDIs focused on issues regarding access to medicines. The IDI guide explored issues regarding existing governance and medicine policy within the NHIS, Supply of medicines (market forces), health information capacity, human resources, health financing and service delivery. All data collection tools were pretested before use in the study.

### Data Analysis

Analysis for quantitative data was done using STATA 11. Frequency and percentages were computed as well as test of association between dependent and independent variables. Chi-square tests were used to determine the test for associations and differences. All tests of significance were carried out at a p value ≤0.05. Data was presented in tables, and narratives as in the result section. For qualitative analysis, interviews were transcribed verbatim. The main themes were identified from responses, coded and analysed using NVivo 11.

### Patient and Public Involvement

Those involved in the research gave verbal and signed consent.

## Results

### Demographic characteristics

296 out of 300 questionnaires were filled correctly. 67.6% were females and 32.4%. Primary beneficiaries comprised 68.2% of the respondents. 72.6% were under paid employment, 9.8% were self-employed and 6.1% were self-employed (see **Table 1)**.

**Table 1:** Demographic characteristics

| Variables | Frequency (%) |
| --- | --- |
| Gender |  |
| Male | 96 (32.4) |
| Female | 200 (67.6) |
| Total | 296 (100.0) |
| Primary Beneficiary |  |
| No | 94 (31.8) |
| Yes | 202 (68.2) |
| Total | 296 (100.0) |
| Employment status |  |
| Unemployed | 18 (6.1) |
| Self employed | 29 (9.8) |
| Paid employment | 215 (72.6) |
| Not applicable | 34 (11.5) |
| Total | 296 (100.0) |

### Accessibility of the scheme

In order to understand the level of access to medicines, respondents were asked questions on the kind of illnesses they had, and the level of access they had to medicines. 89.5% of respondents, said they had acute health issues. 94.9% of respondents sought medical help. 78.4% of the [questionnaire] respondents indicated that the scheme improved their access to care (see **Table 2**).

**Table 2:** The kind of illness respondents sought care for and whether the scheme has improved their level of access

| Variables | Freq. (%) |
| --- | --- |
| **Kind of illness** |  |
| Acute illness | 265 (89.5) |
| Chronic illness | 10.5 (100.0) |
| Total | 31 (296) |
| **Improved access to medicine** |  |
| Yes | 232 (78.4) |
| No | 55 (18.6) |
| Don't know | 9 (3.0) |
| Total | 296 (100.0) |

For the qualitative, one of the IDI respondents believed that all “enrolees had good access to medicines. A few responses of IDI respondents regarding level of access showed are as follows; “access is above average” [ETH]; *very good. You know many people are yet to understand the scheme but for the little I have worked here the scheme is ok*.*”* [*SL*]. There was no discrepancy in access among socio-economic groups. NHIS was reported to have improved access to medicine over the years; *“it has actually given room for people that cannot afford health care to have access to health care. It has actually tried to improve at least marginally it has increased it a little bit. Some people usually just take anything from the patent medicine vendors but now they have access medication without paying. It has actually improved access*.*”*[*SL*].

### Affordability of the scheme

The study found that 82.4% of respondents indicated that costs were partly covered (See **Table 3**). However, 72.9% out of the 82.4% were under paid employment. The remaining 9.5% were either self-employed or unemployed. Socio-economically, among the 82.4% that indicated that costs were partly covered, 31.1% were among the poorest and others, 25.4, 21.1% and 21.7% for the least poor, poor and very poor respectively. 82.8% of respondents indicated that medicines for treatment of common medical conditions are affordable for those with low-income (see **Table 3**).

**Table 3:**
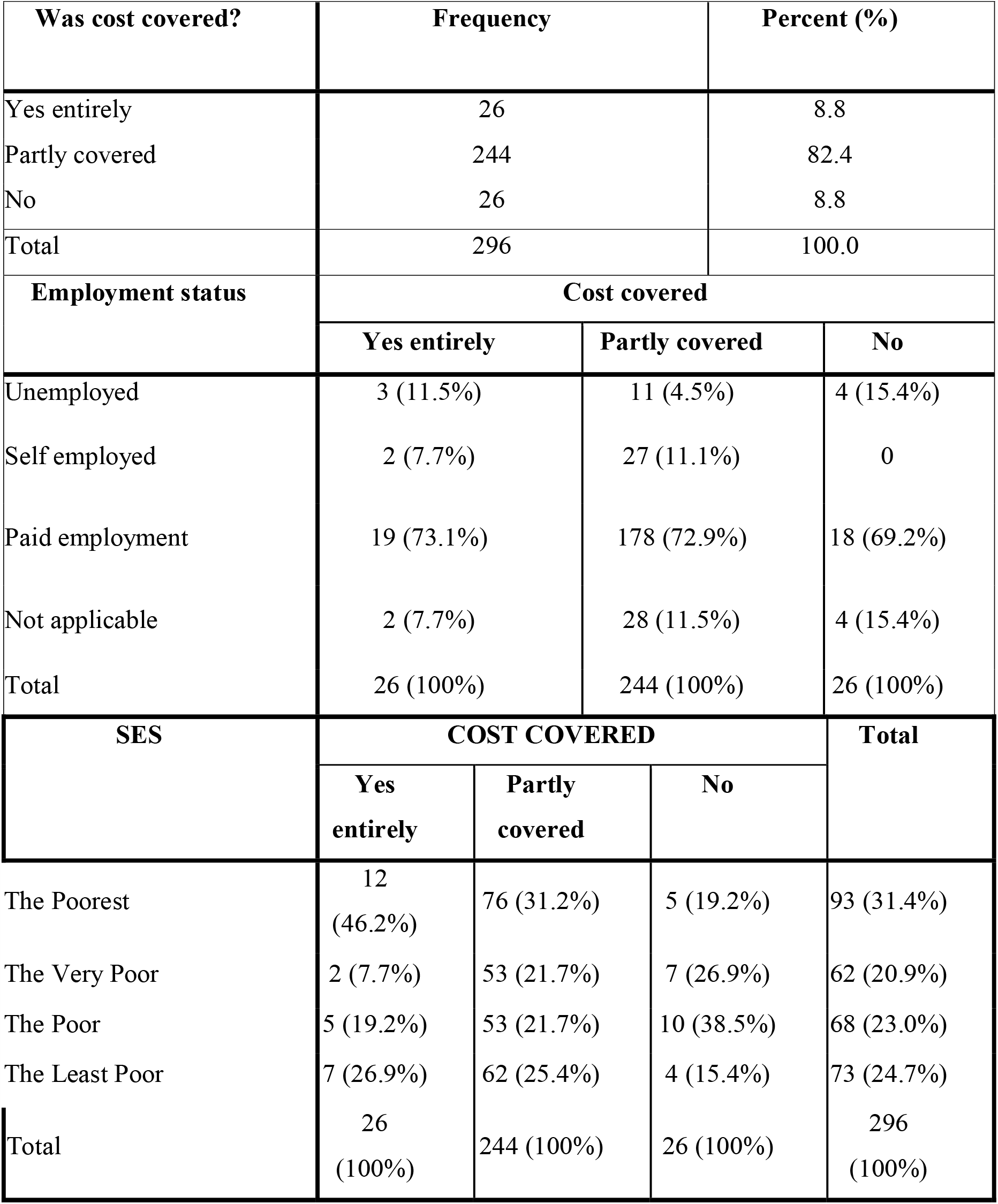
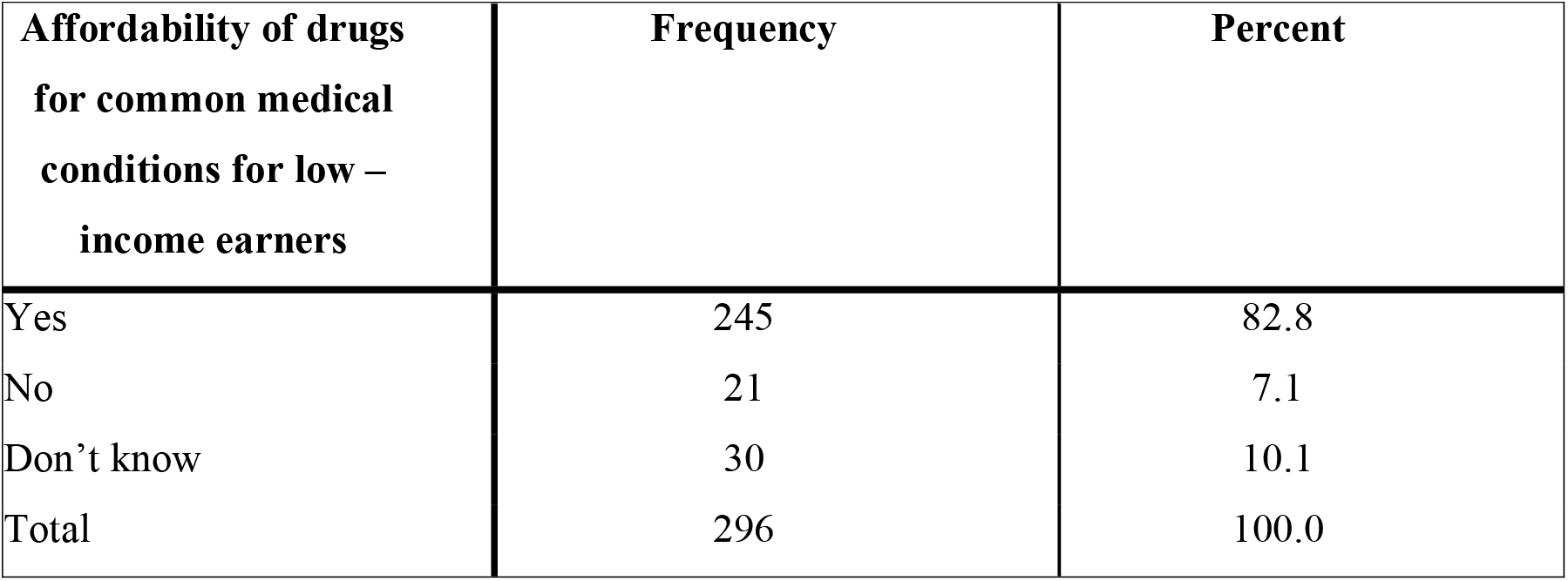
Was cost covered?

According to data collected from IDI respondents, Majority of the respondents opined that NHIS which was designed to subsidize cost of services for users of healthcare is making the private institutions run at loss; *the NHIS is structured in a way that the private institutions are at a loss as compared to the government institutions, because the government institutions have subvention from the government, the government takes care of their overhead while the private work out what they use. So they are already skewed*.*”*[*SL*]. *The price list of drugs presently is not anything to write home about because the NHIS price is very much lower than what is obtainable in the market. These prices are detrimental to the finance of the institution, in other words if we continue, it is a way of running the hospital down and the policy cannot survive for a long time because a lot of private hospitals will opt out”*[*ETH*].

### Availability of the scheme

78% of the respondents were of the opinion that the medicines on the scheme were of good quality (see **Table 4**). This is equally supported by findings from the IDI: “*The medicines provided are good ones”*[*AMH*]. *“They give quality medicine. They give the best within the allocated funding”*[*RC*]. Also, 83.8% indicated the drugs are effective (see **Table 4**). These drugs were not always available. When asked, 47.6% of the respondents were of the opinion that they were not always available, while 45.6% of them believed that the drugs were always available (see **Table 4**). An IDI respondent believes that*”it depends on the health care provider. I can rate it between 50 and 60% depending on the provider but it’s an individual access”*[*SL*].

**Table 4:**
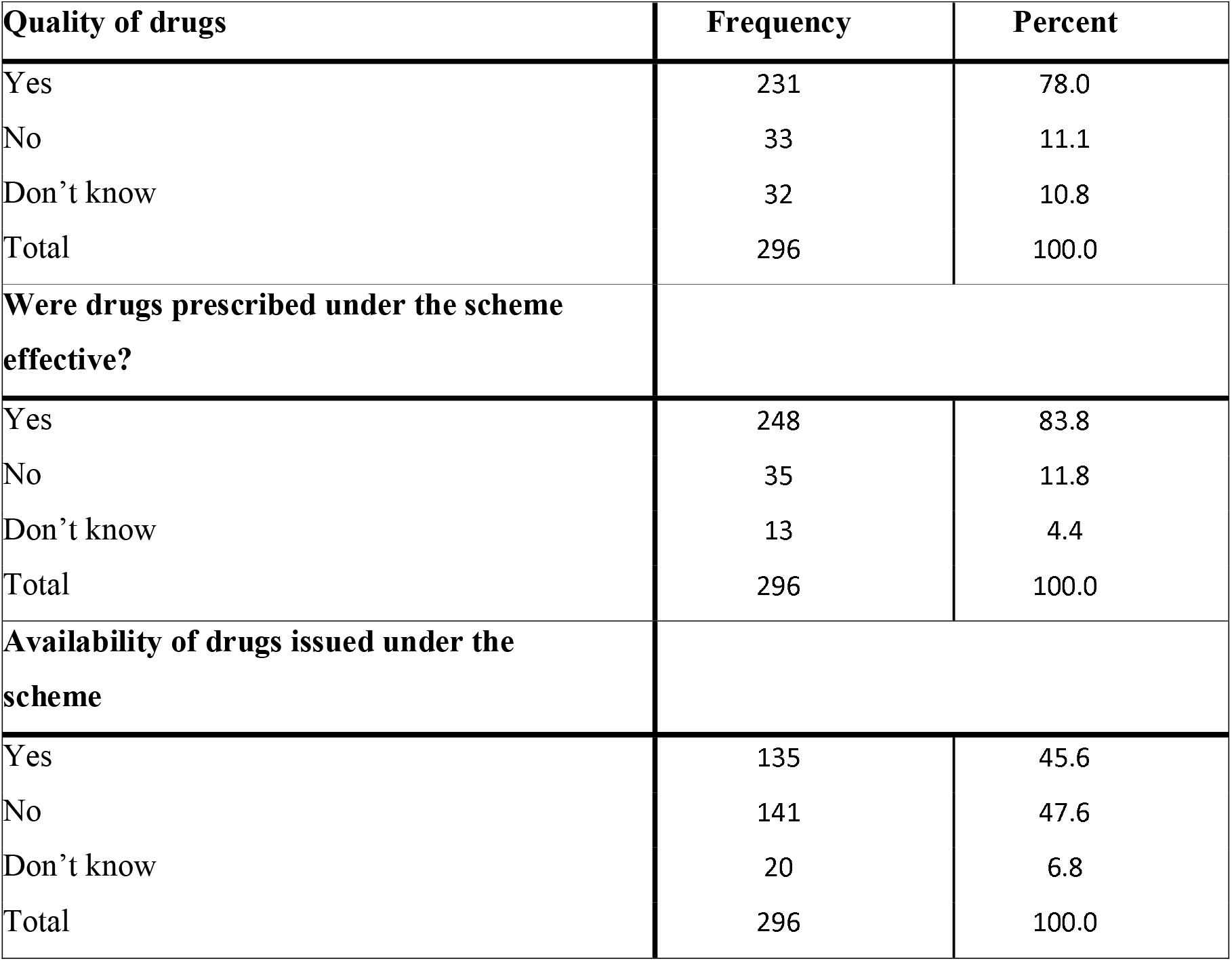
Drugs from the scheme

Staff availability for NHIS facilities were ranked low because in facilities surveyed, staff were not readily available to attend to clients. Results showed that 65.5% indicated that they had to wait long before receiving treatment (see **Table 5**). On the issue of difficulty in getting medicine, 54.4% never had difficulty getting medicine, while 42.6% encountered some difficulties getting medicine (see **Table 5**). Respondents were asked if locally made drugs were more available on the scheme than imported medicine, 49.3% didn’t know, 42.6% indicated “yes” and 8.1% indicated “no” (see **Table 5**).

**Table 5:** Staff availability

| Do you wait long before receiving treatment? | Frequency | Percent |
| --- | --- | --- |
| Yes | 194 | 65.5 |
| No | 93 | 31.4 |
| Don't know | 9 | 3.0 |
| Total | 296 | 100.0 |
| **Did you have difficulty in getting medicine** |  |  |
| Yes | 126 | 42.6 |
| No | 161 | 54.4 |
| Don't know | 9 | 3.0 |
| Total | 296 | 100.0 |
| **Are locally manufactured drugs more available on the scheme than imported drugs?** |  |  |
| Yes | 126 | 42.6 |
| No | 24 | 8.1 |
| Don't know | 146 | 49.3 |
| Total | 296 | 100.0 |

On staff availability, majority of the IDI respondents stated that many of the staff in NHIS accredited facilities are not educated on what is expected or required of them with regards to the scheme. They have very little knowledge on how to run the scheme. This is evidenced by the statement below: “*The desk officers are not even trained. The health care providers don’t even know what the scheme is all about*… *When you go to a hospital that is under NHIS, most times the staff don’t know what the scheme is all about”*.

## Discussion

NHIS was established with the aim to reduce any negative effects of user fees and also to help towards subsidizing the high health care expenses (15). This study sought to assess contextual nature of NHIS with emphasis/focus on accessibility, affordability and availability (3As) and how focus on these three can make NHIS into a more beneficial and long-term scheme towards improving access to medicine.

The constraints to accessing medicines (from the supply side) focuses on healthcare service provider/facilities (16). Among these constraints are facility/service location (17). The scheme, has as one of its objectives, to ensure adequate distribution of health facilities within the federation (18). This is far from realized. The implication of this to the accessibility of the scheme, is that no matter the success recorded now by the scheme, it is still an indication of how much more that needs to be done, especially in the area of accessibility. In order to tackle this issue of accessibility, more facilities should be built proportionately and these facilities across the globe, should be well equipped to handle at the very least, essential medical services. Equally, patients should be able to access the full benefits of the scheme from wherever they are within the country (irrespective of enrolment centre). Accessibility of the scheme cannot be complete without the private hospitals buying in fully into the scheme. The private hospitals are profit minded and to get them to truly buy in, their profit thirsts will have to be quenched.

The NHIS has improved service utilization, this finding is in congruence with other studies (19,20). However, this improvement in utilization tilts a great deal towards salaried workers of all cadre. Although it is of more benefit to low income workers. Low income workers who would have otherwise not been able to have the funds to utilize essential medical services, through the scheme, can now utilize said services. However, the percentage that fall under this group (salaried workers) are nothing compared to a vast majority of unemployed citizens living across the country, who are not able to utilize or have access to essential medical services. More needs to be done in a bid to further improve access to medicine through the scheme. NHIS was designed to cover part costs (10%) for services. Due to the rising cost of health care in the country, the NHIS sought primarily to create a means through which health care can be affordable to all (20). Thus making user fees affordable for enrollees. The 10% co-payment paid by enrollees is the individual’s commitment to the scheme. This creates opportunities for low income earners to afford health care. However, since the scheme only covers salaried workers, the unemployed (those without any means of livelihood) are still left to cater for the full cost of medical services unless they have a family member who enrolls them under their own package. Despite how long the scheme has existed, it’s yet to go beyond the formal sector to cover those at the community level. Total access to essential medicine is still beyond reach.

Availability was another subject raised during the course of this study. Availability of drugs and staff, are key factors in ensuring utilization and access to medicine (21). The drugs provided by the scheme, though of good quality, were not always available. Often times, patients/clients were sent out to the drug stores outside the facility to buy needed drugs. This sometimes meant that they sometimes had difficulty getting medicine. Which could prove a problem for patients in dire need of drugs. This challenge can be taken care by using local pharmacies to dispense drugs free of charge, against the voucher issued to the patient by the doctor. This local pharmacy, can then be reimbursed by the authorities (22). In addition to the drugs, staff were known not to be readily available. This meant that enrollees had to wait long before receiving treatment and this can often prove catastrophic for patients. Depending on the emergency of their health problems, these enrollees may grow inpatient and may decide to use whatever funds they have left to seek medical care. For many of the facilities, even the available staff has no clue as to what is expected of them. In other words, they lacked adequate training needed in performance of their duties. Thus, in a manner of speaking, affecting their relationship with clients and by extension, to access to essential medicine. Therefore, if a patient suffers from a certain ailment, such patient should be able to obtain the right treatment at first call. However, seeing as staff is not always available, this will most likely not be the case (16).

NHIS has as its main objective achieving equitable access to health care in Nigeria (20). They key to achieving this, is a focus on the 3As that matter most - accessibility, affordability and availability (16). The results of this study has already shown what focus on these three can achieve. So far, presence of the scheme alone has been reported to increase utilization and made it a bit more affordable. However, this is not enough as majority of the country’s population are unemployed and are not part of anyone’s benefit package. In line with this, studies (23,24) have suggested that the organization and structure of the bio medical healthcare system of Nigeria, seems to lack in some of those basic components that could enhance access to healthcare. Obuaku (16) pointed out that there is inadequate access to health care services among a large percentage of the population and despite the reforms that has been made by the government, majority of the public health facilities are still short-staffed, ill-equipped, and low on medicines, vaccines and treatments. It has been almost two decades since the introduction of the NHIS and the number of Nigerians covered by the scheme is less than 5% of the population and these are mostly civil servants and corporate workers (and their relatives) in the private sector (25) and those without insurance, who are in need of financial risk protection (more than 90%), are expected to pay out of pocket (26).

Although the scheme has increased/improved utilization, by making the scheme a bit more affordable. There still remains a lot of challenges. These challenges with NHIS from the perspective of the 3As as found by this study, revolves mostly around access to drugs and availability of drugs. Especially regarding private hospitals. The low drug price, low service charge and the fact that they don’t get to go by their own service charge but by that of the NHIS is considered a problem by some of the private hospitals. They believe that they are not making as much in profit as they should: instead of the hospital to be growing financially, it will be running down. If this continues, many hospitals might opt out. Also, there are cases where some of the enrollees develop resistance towards the drugs thus rendering the drugs ineffective. Equally, many of the drugs made available by the scheme have become adulterated. When these clients take it, they don’t actually have the desired effect. Because of it, patients tend to lose confidence in some of these orthodox medicine. There is also, the mentality that anything from overseas is better than what we produced locally. Some prefer the drugs on the high amount than the lower amount because they feel it is more genuine. These information comes from the supply side.

Inadequate availability can be a deterrent to the accessibility of health care. If adequate facilities, skilled staff and (quality) drugs are available and accessible but not affordable, the health services might not be used. This shows the linkage between the 3As. According to World Bank (2019), a great number of Nigeria’s population still live in poverty, without adequate access to basic services, and could benefit from more inclusive development policies. Thus, affordability, in a nation like Nigeria, may be the link that holds all three together and may well be a “golden parachute” towards universal coverage. If people find health services affordable, the number of those seeking healthcare will increase, thus creating a stronger need for health services to be made available and accessible. In all, improved access. However, poor funding of healthcare in Nigeria has been a major barrier to the quality of healthcare service delivery in recent times (26). Thus, the high burden of the costs for healthcare is being borne by individuals and households, which made Nigeria rank as the country with the second highest level of out-of-pocket spending on health in the world (26). There could be a connection between the implementation of NHIS and corruption because the money meant to boost the health sector, most often, ends up in private pockets, which then results to inadequate funding to execute the programme effectively (27). Therefore, availability, accessibility and affordability are structures that can only thrive if adequate funding is released to that effect.

### Study limitations

The study only focused on the health insurance scheme provided by the government. It however, did not look at other health insurance schemes that are already in play or could be in play.

### Conclusion

NHIS has been reported (20,28) to contribute to an increase in health care utilization. However, the scheme is without its perks. A focus on the 3As (accessibility, affordability and availability) for the scheme means that on account of either of the three, all facility categories (private and public) and their interests (where necessary) must be considered in further planning of the scheme to ensure that things hold up fine. In other to ensure a universal coverage, all health care providers (be it private or public) be addressed from their own standpoints - private as private and public as public. Not as one.

Finally, the 3As become a focus when the governing body of the scheme take complete charge of NHIS units in hospitals (private or public) as separate departments or liaison unit, having government employed staff (doctors, nurses, labs, equipment, attendants etc) in all these units across all facilities (private or public). For the private facilities, these staff will be answerable to and paid by the government and not the facilities they operate from. Also, quality drugs and equipment put in use in these units, in all accredited facilities across the globe. Lastly, these staff will be properly trained for the tasks they are employed to carry out.

## Data Availability

All data is available from the lead author and the corresponding author.

## Acknowledgement

Not applicable

## Ethics approval

This study was reviewed and approved by the Health Research Ethics Committee of the University of Nigeria Teaching Hospital, Ituku-ozalla, Enugu state, Nigeria with ID number NHREC/05/01/2008B-FWA00002458-1RB00002323

## Data sharing

Technical appendix, statistical code, and dataset available from the lead author and the corresponding author.

## Informed consent

Written informed consent was obtained from all participants through signatures, thumb prints or verbally. An information sheet, detailing the purpose of the study, proposed participants, and the rights of participants, was given to the participants and reiterated verbally by the researchers.

## Author contributions

### Conceptualization

[Uguru Nkolika Pamela, Ibe Ogochukwu],

### Methodology

[Uguru Nkolika Pamela, Ibe, Ogochukwu],

### Formal analysis and investigation

[Uguru Nkolika Pamela, Ogu Udochukwu Ugochukwu],

### Writing-original draft preparation

[Uguru Nkolika Pamela, Ogu Udochukwu Ugochukwu, Uguru Chibuzo],

### Writing - review and editing

[Uguru Nkolika Pamela, Ogu Udochukwu Ugochukwu, Uguru Chibuzo, Ibe Ogochukwu],

### Resources

[Uguru Nkolika Pamela, Ogu Udochukwu].

